# Coaching for quality improvement under performance-based contracting: a qualitative theory-of-change evaluation in Honduras

**DOI:** 10.64898/2026.05.21.26353487

**Authors:** Wolfgang Munar, L. Esther Aranda, Molly E. Lauria, Pedro Bernal, Cinzia Innocenti, Marvin Rodríguez, María Elena Banegas

**Affiliations:** Department of Global Health, Milken Institute School of Public Health, The George Washington University, Washington, DC, USA; Independent Researcher, Washington, DC, USA; Health, Nutrition and Population Division, Inter-American Development Bank, Washington, DC, USA; Independent Researcher, San Salvador, El Salvador; Independent Researcher, San José, Costa Rica; Independent Consultant, Tegucigalpa, Honduras

**Keywords:** quality management, evaluation, accountability, primary health care

## Abstract

**Background:** Practice coaching is increasingly used to strengthen quality improvement (QI) capacity in primary healthcare (PHC) systems in low- and middle-income countries (LMICs), yet the causal pathways through which it shifts provider behaviour and the systemic conditions that enable or constrain them remain under-theorized. Using a theory-driven qualitative evaluation, we examined how and why a practice-coaching intervention influenced QI in cervical cancer screening (CCS) and antenatal care (ANC) within Honduras’s decentralized PHC system during the third phase of the Salud Mesoamerica Initiative (SMI).

**Methods:** We conducted a within-case explanatory case study. A programme theory was reconstructed before data collection and iteratively refined against evidence drawn from semi-structured interviews with 11 mid-level managers, 6 PHC team medical leads and 2 regional managers, complemented by direct observation and document review. Analysis combined deductive-inductive coding, thematic analysis and pattern matching, with reporting per COREQ.

**Results:** We identified four causal patterns that refined the initial programme theory. Three pathways were activated: novel professional identity among participating managers; data-driven learning and collective efficacy, triggered by verifiable progress on CCS but not for ANC, where outcomes were less attributable to teams’ actions and relational coordination, psychological safety and trust, which provided the interpersonal basis for the first two. A fourth unanticipated pattern showed structural misalignment between coaching’s enabling, learning-based logic and the directive, punitive logic of Honduras’s performance-based contracting environment, limiting gains to localized ‘enabling bubbles.’

**Conclusions:** Coaching can activate meaningful QI pathways in LMIC primary care, but sustained, equitable impact requires purposive alignment between coaching’s learning-oriented principles and the institutional performance-management architecture, and matching of coaching investment to clinical processes with observable, attributable outcomes.

## INTRODUCTION

Expanding access to PHC in LMICs has been a major achievement of global health investment over the past two decades, yet suboptimal provider practices remain a leading cause of preventable mortality and morbidity, thereby limiting effective population-level coverage (1). The Lancet Global Health Commission on High-Quality Health Systems estimated that poor-quality care accounts for a substantial share of excess mortality in low-income settings and called for system-level strategies that go beyond access (2). Over the past four decades, LMIC health systems have increasingly adopted performance management (PM) strategies to align provider behaviour with clinical standards and organizational priorities. However, an evidence gap map (3) of such strategies in LMIC primary care found that the literature is dominated by individual-level financial incentive schemes such as pay-for-performance and results-based financing, while organizationally focused strategies and outcomes remain understudied and poorly theorized.

Practice coaching has emerged as one such organizationally focused strategy. Defined as a collaborative, relationship-based intervention, it is designed to build organizational capacity for QI by empowering frontline teams to use data to make decisions and solve problems (4). Systematic reviews show that coaching can improve clinical processes and, in some instances, patient outcomes, but the evidence base remains concentrated in high-income settings and effectiveness varies substantially across contexts (4–6). Supportive performance-improvement interventions, including supervision, mentoring, and coaching, have been implemented in LMICs (7), but evaluations rarely test explicit causal hypotheses about how and why coaching produces change (8). This theoretical thinness makes it difficult to distinguish between theory failure, where the intervention’s causal logic is itself flawed, and implementation failure, where the logic is sound, but execution was inadequate (9).

### Conceptual framework

We adopt a complexity-informed conceptual framework (10) as our organizing lens for understanding how PM operates within complex health systems. The framework distinguishes two ideal-typical orientations to PM. Directive approaches seek to control provider behaviour through hierarchical answerability, targets, routine monitoring, and extrinsic incentives. Rooted in agency theory (11,12), such approaches assume that providers are self-interested actors whose behaviour diverges from organizational goals unless externally monitored and rewarded; such is the logic underpinning pay-for-performance schemes, whose results in LMICs have been mixed (13). Enabling approaches, by contrast, aim to promote collective responsibility, intrinsic motivation, trust, and team self-organization through team self-organization and data-driven iterative learning. The latter draws on stewardship theory (14) and the premise that providers are intrinsically motivated professionals broadly aligned with organizational goals (15).

The directive–enabling distinction is analytic rather than descriptive: real-world health systems contain both elements, and their interaction shapes how PM strategies evolve. Practice coaching sits within the enabling orientation; collaborative, relationship-based, and designed to strengthen frontline teams’ capacity to identify problems, interpret data, and develop locally owned solutions that do not necessarily enforce compliance with externally imposed targets (7). What remains less well understood is what happens when an enabling intervention is introduced into a PHC system whose context reflects a different, or mixed, set of management logics. The latter is the focus of the study presented here.

### Context and intervention

Honduras is one of Latin America’s poorest and most unequal countries. By 2024, 49.3% of the population lived below US$6.85 per day (2017 PPP), exceeding the Central American average of 37.5%; rural poverty reached 63%, with half of rural inhabitants in extreme poverty (16) (17). These conditions shape both the demand for and the delivery of PHC, particularly in the remote municipalities where the intervention under study was implemented.

Honduras’s PHC system combines decentralization with New Public Management principles, including performance-based contracts and pay-for-performance incentives tied to maternal and child health targets (18–20). The Ministry of Health (Secretaría de Salud - SESAL) retains central stewardship, setting policy, establishing clinical standards, and allocating resources, while delegating service delivery to a diverse set of PHC operators: municipal governments, associations of municipalities, non-governmental organizations, and SESAL-run facilities. SESAL delegated oversight and technical coordination of operators to its regional offices (21). Operators receive flexible budgets tied to maternal and child health targets, with financial penalties for unmet performance. This contractual architecture corresponds to a directive PM environment. The diversity of operator types functioning under distinct institutional arrangements within a single national programme creates natural internal variation, making Honduras an instructive setting for evaluative research on PM strategies such as coaching.

The SMI was a multi-phase, multi-country public–private partnership started in 2011. It was established to improve health system performance and reproductive, maternal, and child health outcomes in the poorest rural municipalities in eight Mesoamerican nations (22). During SMI’s third phase (2019–2022) and extending into one additional year, a practice coaching intervention was introduced in Honduras to strengthen QI capacity of mid-level managers and frontline teams among participating operators. An impact evaluation 02/07/2026 10:22:00 found a positive effect on cervical cancer screening (CCS) and improved QI knowledge among managers, but no significant effect on antenatal care (ANC) guideline adherence and no measurable change in organizational culture.

The evaluation presented here was commissioned after the impact evaluation concluded to help explain how and why the intervention produced its asymmetric effects. It addresses the overarching learning question: How does a practice coaching intervention influence QI in the Honduran PHC system? It is operationalized through three specific questions: (1) To what extent were the hypothesized causal links between coaching activities and the intermediate outcomes of provider capability, opportunity, and motivation realized? (2) To what extent were the underlying causal-link assumptions supported by the empirical evidence? (3) How did the structural diversity of Honduras’s decentralization context shape the activation of the coaching theory?

## METHODS

### Evaluation design

We conducted a qualitative theory-of-change (ToC) evaluation as a within-case explanatory case study of the coaching intervention implemented during SMI Phase 3 in Honduras. ToC evaluation, a type of theory-driven evaluation, treats a programme’s articulated theory as a set of testable causal hypotheses and assesses them against empirical evidence, asking how and under what conditions causal pathways are activated, partially realized, or constrained rather than only whether the programme worked. ToC evaluation is suited to addressing the gaps described earlier. Making causal pathways and programme actors’ assumptions explicit enables researchers to move beyond asking whether a programme worked to explaining how and why its effects did or did not occur (23,24).

The choice of a within-case design was appropriate because the evaluation focuses on a single bounded case: the coaching intervention embedded in Honduras’s decentralized PHC system during SMI Phase 3, allowing us to capture how frontline teams and mid-level managers collectively navigated the overarching national performance-management architecture. The unit of analysis is the coaching intervention as embedded in the Honduran PHC system during SMI Phase 3.

This study operates from a post-positivist integration of scientific realism and pragmatism and is complemented by complexity science perspectives to help understand how program resources trigger psychological and social mechanisms in specific contexts (25–27). It assumes that real causal processes exist within complex systems and can be identified through systematic, theory-driven inquiry, while acknowledging that knowledge of those processes is constructed through both participatory dialogue and empirical testing (28). Accordingly, causal-link assumptions embedded in the programme theory are treated as testable hypotheses rather than fixed truths, and the evaluation seeks coherence between observed evidence and a broader body of theory.

### Programme theory reconstruction

Prior to primary data collection, we reconstructed the programme theory following procedures for surfacing programme logic (29), drawing on programme design and policy documents, participatory workshops with stakeholders, semi-structured interviews with programme actors at multiple system levels, and a targeted scoping of social and behavioural science literature. The reconstructed theory was supported by self-determination theory (30) and adult learning theory (31) at the individual level; social cognitive theory (32) at the interpersonal level; and situated learning theory (33) at the organizational level. Theory reconstruction was also informed by the COM-B model of behaviour change (34) and a systematic process of assumption surfacing (35). The theory was refined after data collection and analysis.

The preliminary programme theory in Figure 1 is represented as a linear causal chain in which coaching activities, including in-person training and field mentoring, would build mid-level managers’ capability (leadership, communication, and QI tool use), which would in turn expand their opportunity to apply new practices (protected time, support to teams, and peer influence), thereby strengthening their motivation (beliefs, goals, and habitual routines). Motivation was hypothesized to translate into two proximal behavioural outcomes: improved managerial supervision quality and sustained team-level QI practices, which would jointly drive the distal service-delivery outcomes of CCS and ANC adherence.

**Figure 1.**
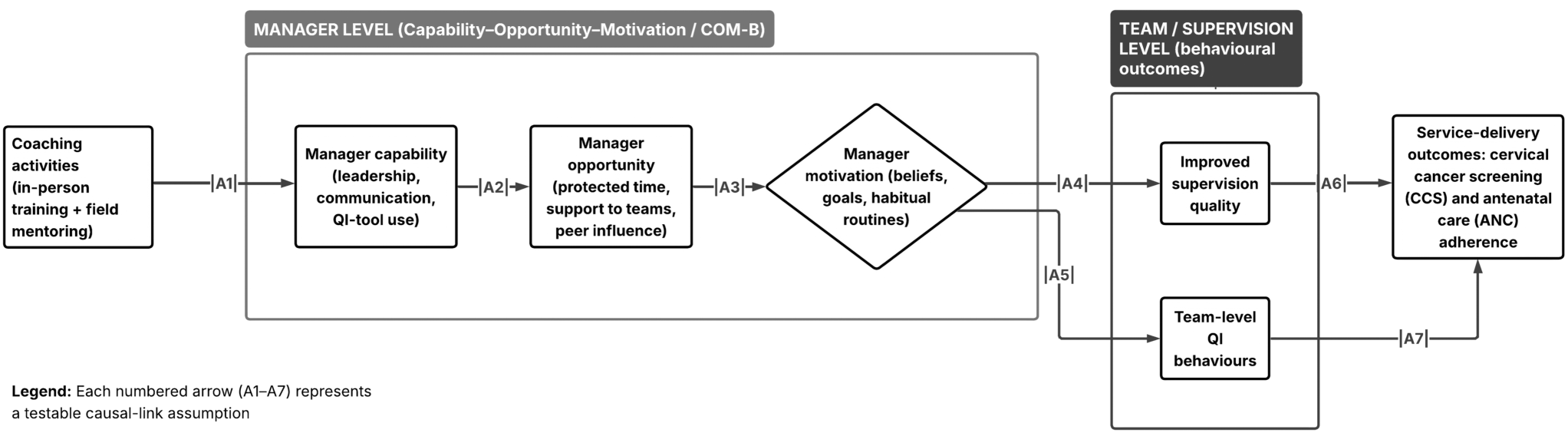
Preliminary Programme Theory.

### Data sources

Primary data were collected through 19 semi-structured interviews collected between March and May 2024 and direct observation of the final coaching training module in November 2023. Participants included mid-level managers, PHC team medical leads, and two of the three regional SESAL directors overseeing the regions in which the intervention was implemented (Table 1). Participants were invited individually via email, and interviews were conducted virtually in Spanish, with each participant interviewed only once. The interviewer was a native Spanish-speaking female senior qualitative researcher who had been involved in prior Phase-2 research of the SMI in Honduras and had no knowledge of the Phase 3 participants. Mean interview duration was 65 minutes; interviews were audio-recorded, with brief field notes taken by the interviewer, and professionally translated into English for analysis; only the interviewer and participant were present during the interviews. Two PHC leads and one SESAL regional director could not be interviewed due to time conflicts, and there were no withdrawals.

**Table 1.** Participant characteristics by respondent type.

| **Respondent type** | **Total** | **Women** | **Men** |
| --- | --- | --- | --- |
| Mid-level managers / supervisors and medical coordinators | 11 | 8 | 3 |
| PHC team medical leads | 6 | 5 | 1 |
| Regional SESAL Managers | 2 | 1 | 1 |
| Total | 19 | 14 | 5 |

Participants were recruited through a purposive, maximum-variation strategy designed to maximize the information power of the sample with respect to the causal hypotheses being studied (36). Recruitment proceeded in two stages. First, at the operator level, we invited mid-level managers from all operators engaged in the intervention, ensuring that the full institutional range of contracted providers was represented. Second, within each Operator, we drew a smaller, nested sub-sample of PHC team medical leads, selected for being supervised by the sampled managers and for their direct, first-hand experience with team-level coaching activities. Finally, we interviewed two regional SESAL directors.

An interview guide (Appendix A) was developed from the programme theory and the above theoretical framework; it covered eight thematic areas, including training history, supervision and monitoring practices, knowledge of national CCS and ANC standards, responses to the coaching intervention, QI implementation experience, and perceptions of external influences on national guidelines implementation. Secondary data included Honduran Government policy documents; sector diagnostics from regional development banks and other multilateral organizations; programme documents and IHME performance verification data sets; and targeted peer-reviewed and grey literature.

### Data analysis

Analysis followed a structured, iterative theory-testing strategy. We built chronological narratives, then three coders applied combined deductive coding derived from the programme theory and theoretical framework, and inductive coding to identify emergent themes, working iteratively among the articulated theory, the impact evaluation findings (37), and the primary qualitative data. Pattern matching (38) was used to compare hypothesized and observed causal patterns, and alternative explanations were systematically surfaced. Coding was conducted in two cycles by a three-member team in ATLAS.ti, with intercoder consistency established on shared transcripts in the two cycles of coding; an audit trail documented coding decisions and the evolution of the analytical framework. The codebook is presented in Appendix A.

Operational and budgetary constraints impeded member checking. Credibility was instead strengthened through triangulation across interviews, programme documents, and the results of the prior impact evaluation, as well as through systematic searches for disconfirming evidence. Dependability was supported by collaborative two-cycle coding and independent coding by team members not previously involved in programme implementation. Reporting follows the COREQ checklist presented in Appendix A.

### Use of artificial intelligence

The authors used Grammarly Pro solely for grammatical review; all analytical interpretations, theoretical claims, and evidentiary judgments are the authors’ own. The authors take full responsibility for the integrity and accuracy of all manuscript content.

## RESULTS

We identified four causal-link patterns through which the coaching intervention interacted with Honduras’s PHC system. Three describe activated pathways linking coaching activities to changes in mid-level managers’ and frontline teams’ capabilities, opportunities, motivation, and practices. A fourth, unanticipated pattern describes the systemic conditions that constrained those pathways and confined their effects to localized “enabling bubbles” of improvement. Table 2 summarises the patterns and their evidential support; Table 3 maps illustrative quotes to each pattern by respondent type. Additional supporting evidence is presented in Appendix B.

**Table 2.** Causal-link patterns, theoretical foundations, and evidential support.

| Pattern | Causal-link assumption tested | Theoretical foundation | Evidential support |
| --- | --- | --- | --- |
| 1. Professional identity transformation | If managers receive integrated soft- and hard-skill training plus mentored field practice, then they will shift from directive to facilitative supervision. | Adult learning theory (Knowles, 1984); self-determination theory (Ryan & Deci, 2000); COM-B model (Michie et al., 2011). | Consistent across mid-level manager interviews; corroborated in part by a PHC team lead; partial train-the-trainer fidelity loss noted (1 case). |
| 2. Collective efficacy and data-driven learning | If teams apply QI tools and receive timely data feedback, then collective efficacy will emerge and behaviour change will be sustained. | Social cognitive theory (Bandura, 2000); situated learning theory (Lave & Wenger, 1991). | Strong reinforcing loop for CCS, where indicators are observable and attributable; weaker/attenuated loop for ANC; loop fragile at the re-measurement step when indicators fell outside teams' data range. |
| 3. Relational coordination, psychological safety, and trust-building | If coaching strengthens communication and supportive leadership, then teams will experience increased psychological safety and improved coordination. | Self-determination theory (Ryan & Deci, 2000); social cognitive theory (Bandura, 2000); situated learning theory (Lave & Wenger, 1991). | Cited across manager and PHC-lead interviews; rapid catalytic shifts triggered by external validation during mentoring visits. |
| 4. Incentive misalignment and systemic constraints | Not in initial theory. Emergent: enabling coaching logic conflicts with the directive, inspection-oriented logic of performance verification, confining gains to localized enabling bubbles. | Performance management in complex adaptive systems (Newton-Lewis et al., 2021); stewardship theory (Davis et al., 1997) vs. agency theory (Eisenhardt, 1989; Mitnick, 1975). | Convergent across PHC managers and team leads (logic clash; punitive audits and fund-withdrawal for missed targets; multi-month payment delays — a distinct mechanism; resource shortages; turnover/contract insecurity). Regional-director accounts provide complementary, theory-consistent context, not convergent confirmation. Suggestive, single-source signal of target-softening from one non-coached PHC lead (HF-04). |

**Table 3.** Illustrative quotes by causal-link pattern and respondent type.

| **Pattern** | **Mid-level manager / Coordinator (OP-)** | **PHC team medical lead / Regional director** |
| --- | --- | --- |
| **1. Identity transformation** | "not accusing or pointing fingers but seeing first what is positive" (OP-12) | "when coaching came ... our chip began to change." (HF-03) |
| **2. Collective efficacy & data-driven learning** | "the measurements, the metrics, and the graphs, seeing where we were and how we're moving forward, how we're achieving our goals — I feel like that's important." (OP-08) | "we stopped seeing that indicator as a cold number and started seeing it more as a person" (HF-03) |
| **3. Psychological safety & trust** | "my team sees me as a co-worker; we are horizontal." (OP-02) | "when the team of international coaching agencies that came in started congratulating us ... they were motivated and totally changed." (HF-03) |
| **4. Incentive misalignment** | "that is punitive for us. If we don't have something, they take away points ... if we don't have it, they take our money." (OP-10) | *For context only:* "if you do not have a goal ... then you are just going to be there and give the minimum you can give." (DIR-02) |

### Pattern 1. Professional identity transformation and enhanced confidence

Mid-level managers consistently reported that their prior clinical education had not prepared them for supervisory roles, leading them to adopt the directive styles prevalent in their environment. One located this in the training of the nursing cadre: “the licensed nurses, we are only made to command … I am the boss. I do this. And I want this and that” (OP-01). Another recalled a harsher prior style: “before, I achieved everything, perhaps in an angry way … in a rough way” (OP-02). Participants framed the coaching’s combination of quality-improvement tools and soft skills as filling a gap left by clinical training and experienced the learning as personal: “I have felt that I have changed personally; … it really makes us see and reflect in a professional and personal way … we must change” (OP-12).

Managers described a shift from enforcer to facilitator. This was articulated as a change in style: “it is not the same for you to tell me ‘do’ as it is for you to say, ‘let’s do’” (OP-01), as a horizontal relationship: “my team sees me as a co-worker; we are horizontal” (OP-02), and as a move away from blame: “not accusing or pointing fingers but seeing first what is positive” (OP-12). Managers linked the new style to greater team ownership, “they feel more responsible for undertaking the commitments” (OP-01), and to improved communication that “gives us better results” (OP-02).

These accounts were corroborated, at least in part, by a PHC team lead who recognized the directive baseline, “before … this has to be done. You have to do this, this, and that’s it,” and a subsequent reorientation they attributed to the intervention: “when coaching came … our chip began to change” (HF-03). The same lead described initial resistance that was later reversed by external validation during the first mentoring visit, an emotional reorientation examined more fully in Pattern 3.

The transition was not uniformly welcomed, suggesting the shift may have been negotiated rather than automatic; one team initially “wanted the boss to be stricter” (OP-01), indicating that enabling supervision may need to incorporate accountability rather than displace it.

### Pattern 2. Data-driven learning and collective efficacy

Managers and team leads described a recurring feedback loop in which baseline measurement, root-cause analysis, and re-measurement made progress visible and, in turn, promoted motivation and continued use of QI tools. One supervisor located the mechanism in the data itself: “the measurements, the metrics, and the graphs, seeing where we were and how we’re moving forward, how we’re achieving our goals — I feel like that’s important” (OP-08). For some, this reframing of measurement was experienced as a shift in how patients themselves were seen: “we stopped seeing that indicator as a cold number and started seeing it more as a person” (HF-03). Teams translated these routines into concrete, verifiable workflows, such as a patient sign-in register confirming “the day and the date that the result was delivered to verify” (HF-01) timely delivery of results and structured weekly review formats that “made it much easier to identify where the problem was” (HF-02).

This feedback loop was topic-general rather than specific to CCS. It also operated on the ANC side, where one manager reported that once “we had positive results, almost immediately… they were also motivated” (OP-06), and a team lead noted that “last year we had a low indicator, this year it has improved a lot. That was my motivation” (HF-06). Where it functioned, visible gains and collective effort reinforced one another: “The whole team was very satisfied. We were all fully involved” (OP-07).

The loop was fragile, depending on data that could be controlled by teams rather than on the clinical topic itself. It failed during the re-measurement step whenever the indicator was outside the team’s data range: private-sector cytology that “we are not going to have them [counted]” (OP-03), denominator mismatches with their annual operational plans, or a mid-stream measurement redefinition that left one team lead unable to “make heads or tails” of a new indicator (HF-05). In such cases, the same mechanism produced demotivation rather than efficacy, a dynamic that connects to the systemic constraints described below in Pattern 4.

### Pattern 3. Relational coordination, psychological safety, and trust-building

The third pattern functioned as the interpersonal connective tissue that allowed the first two to take hold. Supervision was increasingly experienced as helpful rather than punitive: one supervisor distinguished her work as enabling: “it is not that we will grade it … we will [find] alternatives” (OP-08), and another described shifting from working “by pressure” to working “by encouragement” (OP-05). Reframed as partners, managers conveyed that “we are all working towards the same goal and that we are together and that they are not alone” (OP-02), so that teams “feel that they are not alone, that they have a backup, that they have the support to improve anything” (OP-02). This relational reframing lowered the threshold for raising problems without blame, reinforcing the shift away from blame described in Pattern 1; team leads corroborated this by describing their supervisor as the one who “supported us” through challenges (HF-06). Some respondents also associated the new climate with a lighter workload, with one supervisor compensating for field overload “with free time” (OP-11). External validation appeared to reinforce the shift; one team lead recalled initial resistance, when staff felt QI projects as “more work,” reversed during the first mentoring visit, “when the team of international coaching agencies that came in started congratulating us … they were motivated and totally changed” (HF-03), an emotional reorientation reiterated by a coordinator who felt such engagement meant “we felt more motivated” (OP-01). Together, these accounts suggest that supportive external feedback and horizontal relational framing provided the trust on which the slower identity transformation of Pattern 1 and the data-driven loop of Pattern 2 depended.

### Pattern 4. Incentive misalignment and systemic constraints

A fourth pattern, unanticipated by the initial programme theory, emerged from descriptions of the institutional environment, in which the coaching’s enabling, learning-based logic met the directive, inspection-oriented logic of SESAL regional offices’ performance verification. Managers across operators drew a sharp distinction: SESAL monitoring mattered “because money is at stake … If they do not comply with the indicators, then they lose points while supervision is enabling” (OP-08), and audits were experienced as unidirectional, conducted by auditors who “leave a lot of commitments here that sometimes people don’t even understand. And they leave and never come back” (OP-10). The same supervisor described verification as “punitive for us. If we don’t have something, they take away points,” adding that “if we don’t have it, they take our money” (OP-10). Where indicator construction diverged from annual operating plans’ targets, the result was demotivation rather than learning: “right now for us it is a big demotivation … most of the teams feel that … no, we are not going to meet the cytologies” (OP-10), addressing the re-measurement failure flagged in Pattern 2. These tensions were compounded by systemic constraints: chronic resource shortages, with one lead noting that “if you are going to do cytology, there are no gloves” (HF-04); multi-month payment delays, where a coordinator reported that the cytology laboratory threatened to stop reading samples because the operator “had not paid in almost five months” (OP-07), and another described recurring “problems with payment” that left staff “unmotivated” (OP-06); and high staff turnover compounded by contract insecurity, with one operator retaining only “three out of the ten doctors we had” (OP-10), and a lead returning to find “half of the staff that had been trained in coaching was no longer there” (HF-06), noting “most of us have a contract signed until June of this year … I feel a little bit uncertain” (HF-06). Within this directive frame, very low screening targets could be met defensively; one non-coached lead recounted a declining CCS target sequence: “First, it was 33, more or less 35. It went down to 23, 17, and now we have 13,” concluding that “since the goal was low, it was not very difficult to achieve” (HF-04). Given that the single source mentioning such target-softening, an argument for gaming cannot be constructed, although the 13-per-month target was corroborated by the corresponding supervisor (OP-03).

Regional directors construed the contextual architecture described by managers and team leads very differently: as legitimate stewardship ensuring “the funds and resources are being invested in the best way” (DIR-01), and as a motivational necessity, since without established goals, a worker “[is] just going to be there and give the minimum you can give” (DIR-02). One regional director even wished monitoring were “a little bit stricter”. This perception gap among regional directors, managers, and frontline providers (Box 1) meant that the demotivators most salient to frontline cadres were largely invisible to the tier verifying their performance, confining quality improvements to localized “enabling bubbles” bounded by SESAL’s pervasive directive logic.

#### Box 1. Two views of the performance monitoring system

**Frontline experience (OP-01):** “*They do something called technical audits where they come, they identify the problem, and they leave a lot of requirements here that sometimes people don’t even understand. And they leave and never come back. They come back one or two years later… There is no review of previous commitments or follow-up of the process.”*

**Regional managerial framing (DIR-02)**: “*With constant monitoring of the health centres, people work a little bit based on a goal… If you do not have a goal to establish, if you do not know your indicators, then you are just going to be there and give the minimum you can give*.”

## DISCUSSION

This theory-of-change evaluation identified four causal-link patterns that, taken together, help refine the intervention’s initial programme theory. Three describe activated pathways at the individual, team, and interpersonal levels: professional identity transformation from directive to supportive supervision (Pattern 1); data-driven learning and collective efficacy, through which teams sustained behaviour change by verifying measurable progress (Pattern 2); and relational coordination, psychological safety, and trust-building, which constituted the interpersonal basis for the first two (Pattern 3). A fourth pattern, unanticipated by the initial theory, describes a structural misalignment between the intervention’s enabling, learning-based logic and the directive, inspection-oriented logic of Honduras’s performance-based contracting environment (Pattern 4). These patterns are synthesized in the refined programme theory in Figure 2. In the refined model, three coaching-activated mechanisms generate localized QI gains within enabling ‘bubbles’, which are constrained by the directive performance-management logic of the wider health system (Pattern 4). By systematically mapping these qualitative configurations, the evaluation provides a plausible framework for distinguishing potential implementation failures from underlying theoretical limitations (9) within the coaching intervention. The evidence suggests that the stagnant outcomes in ANC guideline adherence did not primarily stem from an implementation failure; informant accounts indicate that the intervention successfully activated its intended relational processes, corresponding to meaningful shifts in managerial identities (Pattern 1) and the cultivation of interpersonal trust (Pattern 3) across both clinical tracks. Instead, the divergence points to a boundary limitation or a localized theory failure within the coaching model’s foundational assumptions. While the initial programme theory presumed that data-driven learning loops would operate uniformly across different clinical domains, the results suggest that because multi-step ANC processes are less immediately observable or attributable to frontline actions, the psychological mechanism linking data to collective efficacy likely faltered, pointing to a breakdown in the behavioral feedback loop. Similarly, the apparent containment of CCS gains within localized ‘enabling bubbles’ suggests that, while the intervention’s core behavioral theory is supported when metrics are highly observable, its macro-level scaling assumptions did not fully anticipate the structural counter-pressures of the surrounding directive performance-management environment. Untangling these dynamics illustrates the utility of ToC evaluations in shifting health systems research away from binary judgments of programmatic success toward a more nuanced mapping of institutional and task boundaries.

**Figure 2.**
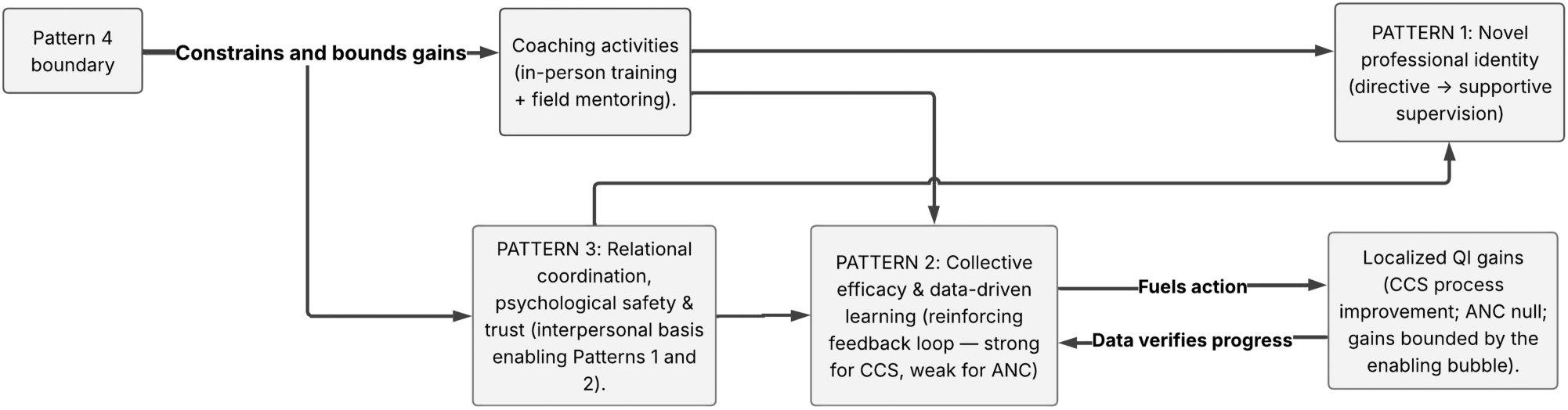
Refined Programme Theory.

The clearest explanatory contribution of this evaluation is its provision of plausible explanations for the asymmetric findings in the impact evaluation that preceded this study. Cervical cancer screening is a service whose process indicators may have made team performance observable, and whose clinical impact is occasionally vivid and proximate. These features appear to have activated the Pattern 2 reinforcing loop: data-verified success strengthened collective efficacy, which motivated continued application of QI tools, which produced further measurable gains, consistent with social cognitive theory’s predictions (Bandura, 2000). Antenatal care adherence, by contrast, is a multi-step process whose outcomes are harder to verify through routine indicators. The reinforcing loop appears to have been weakly activated, providing a plausible mechanism-level explanation for the null ANC effect that an outcome-focused study cannot supply on its own. Consistent with Pattern 2, the loop also faltered at the re-measurement step whenever the relevant indicator lay outside the team’s data range, for example, private-sector cytology that teams could not count or denominators misaligned with the operational plans, so that the same measurement mechanism that drove efficacy elsewhere instead produced demotivation, linking this fragility directly to the systemic constraints of Pattern 4. Also, provider adherence, while necessary, is not sufficient to address the demand side of ANC and CCS. This may generalize beyond Honduras: coaching interventions may produce stronger, faster gains where clinical processes generate observable, attributable, and frequent feedback, and weaker gains where they do not.

The statistically significant CCS findings from the SMI evaluation require qualification. Our evaluation assessed a programme theory rather than a counterfactual, supporting the inference that coaching may have contributed to observed CCS improvements, but alternative explanations are possible. In particular, very low baseline performance targets and punitive verification may have fostered defensive reporting behaviours. Several participants described accountability as sanction-oriented. The narrative of a declining cytology target raises the possibility that some CCS gains reflect strategic target softening rather than improved practice, particularly where small improvements shifted indicators above the minimal thresholds. This aligns with concerns in the literature that performance verification can distort effort and inflate reported performance (39). However, this inference rests on a single respondent; while the target level itself was corroborated by the corresponding supervisor, the gaming interpretation was not. We therefore do not assert that gaming explains the CCS result and treat it as suggestive only. Acknowledging this alternative explanation is nonetheless necessary to qualify the CCS-positive inference and frame the comparison with ANC outcomes.

Interpreting the null findings for ANC guideline adherence requires more than just supply-side explanations. While our refined theory suggests that supply-side issues, such as the visibility of indicators and immediate feedback, play a role, they don’t fully account for the findings. In rural settings, demand-side barriers such as transport costs, travel distances, and language gaps also limit ANC uptake, beyond what coaching can address. These are structural, socio-economic factors outside the scope of frontline interventions. Our model focuses on service delivery, highlighting supply-side pathways, but overlooks demand-side constraints that significantly impact accessibility. Therefore, when comparing ANC and CCS, the differences lie not only in how their performance signals are observed but also in service accessibility. Recognizing this demand-side dimension clarifies the boundaries within which coaching can influence outcomes.

Our findings indicate that coaching’s enabling logic, triggered by collaborative problem-solving, data for learning, and supportive supervision, operated within an institutional context characterized by a directive logic: performance-based contracts and pay-for-performance with explicit financial penalties, episodic technical audits experienced as unidirectional and demoralizing, and indicator monitoring tied to the withdrawal of funds for missed targets, a sanctioning mechanism distinct from the multi-month delays in routine payments to operators that providers also described. The PM framework that guided our theoretical model (10) anticipated that such mixed institutional environments would produce outcomes shaped by the interaction of the two logics rather than by the PM intervention alone, and our evidence seems to confirm such a prediction in behavioural terms. The two logics did not simply coexist; they generated competing demands on frontline managers and teams, who were asked to conduct QI routines while being audited for compliance with indicators. They also triggered the experiential demotivators most significant to providers: delayed payments, supply shortages, and job insecurity arising from short-term contracts and turnover, which regional managers overseeing the system largely disregard. The result was an ‘enabling bubble’ phenomenon (40): real but spatially and temporally bounded improvement, contingent on local relational conditions and vulnerable to system constraints. This finding is consistent with the conceptualization of health systems software and hardware (41,42); with Hood’s (43) caution that performance-by-numbers regimes can simultaneously enhance and obstruct service performance, depending on organizational culture and the credibility of metrics; and with realist evaluation evidence from other LMIC settings (44–48) that motivational mechanisms are highly context-contingent.

Three implications for policy and practice are outlined below. First, sustaining coaching’s gains in LMIC PHC systems requires deliberate alignment between the intervention’s enabling principles and the institutional performance-management architecture, rather than co-location alone. Design considerations include remaking regional performance verification engagement as developmental rather than punitive, decoupling fund-withdrawal triggers from indicators where coaching for QI is in active use, and creating shared dashboards that make frontline experiential data legible to regional managers, which we term supportive accountability. Embedding such design changes requires engaging macro-level decision-makers within SESAL: without national-level ownership, alignment efforts risk remaining regional and temporary rather than institutionalized and sustained beyond the life of donor-funded coaching. Second, programme designers should match coaching investment to the feedback observability of the targeted clinical processes; where outcomes are diffuse, complementary measurement infrastructure (proxy process indicators, structured patient-flow tracking) is likely a precondition for the Pattern 2 loop to activate. Third, the train-the-trainer fidelity weakness that surfaced in Pattern 1 suggests that scale-up should preserve field mentoring rather than rely solely on train-the-trainer, cascade approaches, even at the cost of slower geographic reach.

Two research priorities emerge. First, longitudinal mixed-methods studies are needed to characterize how directive and enabling performance-management logics interact over time within single decentralized health systems, and whether “enabling bubbles” expand, persist, or collapse once external coaching support is withdrawn. Also, comparative work across SMI countries would test whether the four causal-link patterns generalize or whether their relative strengths vary with institutional configurations. Such work would inform the design and evaluation of coaching interventions in decentralized health systems and clarify the conditions under which enabling interventions can reshape, rather than merely coexist with, directive performance-management environments.

### Strengths and limitations

Study strengths include the explicit theory-testing design, the iterative theory refinement, the triangulation of interview data with direct observation and programme documentation, and the pairing with a prior impact evaluation. Limitations should be acknowledged. Sampling was limited to managers, PHC team leads, and regional directors; neither other team members nor patients were interviewed, and their perspectives may modify Pattern 3. The evaluation took place at the end of an established multi-phase programme, and even if the intervention included all operators in SMI areas, it raises the possibility that the most successful PHC teams were over-represented. Relatedly, although we interviewed two of the three regional directors overseeing the intervention, the regional-tier evidence informing Pattern 4 and Box 1 should be read as theory-consistent, whereas the manager- and provider-level evidence underpinning Patterns 1–3 is substantially more convergent.

## Conclusion

Practice coaching can activate meaningful causal pathways toward quality improvement in PHC systems of LMICs, transforming supervision from directive to supportive, catalyzing data-driven collective learning, and building the relational conditions on which both depend. In Honduras, however, these gains remained confined to localized “enabling bubbles” because the intervention’s enabling logic operated within an institutional architecture organized around a directive form of PM. Coaching’s effects were also asymmetric across clinical processes, emerging strongly for CCS, where progress was observable and attributable, but not for ANC, suggesting that coaching investment should be matched to processes that generate observable, attributable feedback. Ultimately, by moving beyond an exclusive focus on counterfactual compliance, this study illustrates that what is often categorized as poor implementation in LMIC health systems may instead reflect a mismatch between the intervention’s underlying explicit or implicit theories of task complexity and macro-institutional alignment. Sustained system-wide impact, therefore, appears to depend on a purposive alignment between coaching’s learning-oriented principles and the broader performance-management architecture, which requires national-level ownership to transition from punitive monitoring to supportive accountability. Sustained system-wide impact, therefore, requires purposive alignment between coaching’s learning-oriented principles and PM system architecture, not only at the regional tier but with national-level ownership, an approach we term supportive accountability. Further longitudinal and comparative research is needed to characterize how directive and enabling PM logics interact over time in decentralized health systems, and to inform the design of coaching interventions that can reshape, rather than merely coexist with, prevailing institutional logics.

## Supporting information

Appendix A - Guidelines, Codebook, COREQ

APPENDIX B: Indicative quotes

## Data Availability

No data are available. The study draws on confidential key-informant interviews conducted with a small, purposively selected sample. The latter and the limited number of identifiable role-holders within the participating organisations make release of the transcripts, even in de-identified form, unsafe. The data are therefore not publicly available, in line with the ethical restrictions approved by the George Washington University IRB (protocol NCR245559) and with the written informed consent obtained from participants. These restrictions cannot be waived.

## Patient and public involvement

Patients and members of the public were not involved in the design, conduct, reporting, or dissemination plans of this research.

## Ethics approval and consent to participate

Ethical approval was obtained from the corresponding author’s institutional review board [name and reference number anonymized for peer review] and from the relevant national health authority in Honduras. All participants provided written informed consent before participating in the study. Participation was voluntary, and participants were informed of their right to withdraw at any time without consequence. Confidentiality and anonymity were assured throughout data analysis and reporting.

## Data availability statement

No data are publicly available. The study draws on confidential key-informant interviews conducted with a small, purposively selected sample of participants. The limited number of identifiable role-holders makes release of the transcripts, even in de-identified form, a risk to participant confidentiality. The data are therefore not publicly available, in line with the ethical restrictions approved by the Institutional Review Board and the terms of the written informed consent obtained from participants. Reasonable, restricted requests from qualified researchers may be directed to the corresponding author and will be considered on a case-by-case basis, subject to ethical and confidentiality safeguards.

## Acknowledgements

The authors thank the mid-level managers and primary healthcare team medical leads who participated in the interviews and candidly shared their experiences. We are grateful to colleagues at [institutional partners anonymized for peer review] for their support. Any errors or interpretive judgements are the authors’ alone.

## Supplementary materials

Appendix A — Interview guidelines, codebook, and COREQ checklist. Appendix B — Indicative quotes by causal-link pattern and respondent type.

## Declarations

### Ethics approval and consent to participate

Ethics approval was obtained from the Institutional Review Board of The George Washington University (IRB #200213) and the National Health Secretariat of Honduras.

### Consent for publication

Consent for publication is not applicable as no individual patient or participant data/images are presented in an identifiable form.

### Availability of data and materials

The qualitative datasets generated and analyzed during the current study are not publicly available due to participant confidentiality and ethical restrictions related to the highly visible professional positions of the key informants (health facility coordinators and regional directors). However, anonymized transcript segments remain available from the corresponding author upon reasonable academic request.

### Authors’ contributions

WM conceptualized the study framework and led the analysis. CI conducted data collection LEA, MEL and MR conducted coding and thematic mapping. WM and LEA led draft preparation. PB led the preceding quantitative evaluation. CI and MEB provided administrative, logistical, and local coordination support. All authors read and approved the final manuscript.

## References

1. Garcia-Elorrio E, Rowe SY, Teijeiro ME, Ciapponi A, Rowe AK. The effectiveness of the quality improvement collaborative strategy in low-and middle-income countries: A systematic review and meta-analysis. Plos One. 2019;14(10):e0221919.

2. Kruk ME, Gage AD, Arsenault C, Jordan K, Leslie HH, Roder-DeWan S, et al. High-quality health systems in the Sustainable Development Goals era: time for a revolution. The Lancet Global Health. 2018;6(11):e1196–252.

3. Munar W, Snilstveit B, Aranda LE, Biswas N, Baffour T, Stevenson J. Evidence gap map of performance measurement and management in primary healthcare systems in low-income and middle-income countries. BMJ Global Health. 2019;4(Suppl 8).

4. Grumbach K, Bainbridge E, Bodenheimer T. Facilitating improvement in primary care: the promise of practice coaching. Issue Brief (Commonw Fund). 2012;15(1):1–14.

5. Walunas TL, Ye J, Bannon J, Wang A, Kho AN, Smith JD. Does coaching matter? Examining the impact of specific practice facilitation strategies on implementation of quality improvement interventions in the Healthy Hearts in the Heartland study. Implementation Science. 2021;16(1).

6. Wang A, Pollack T, Kadziel LA, Ross SM, McHugh M, Jordan N. Impact of practice facilitation in primary care on chronic disease care processes and outcomes: a systematic review. J Gen Intern Med. 2018;33(11):1968–77.

7. Vasan A, Mabey DC, Chaudhri S, Brown Epstein HA, Lawn SD. Support and performance improvement for primary health care workers in low-and middle-income countries: a scoping review of intervention design and methods. Health policy and planning. 2017;32(3):437–52.

8. Floate H, Durham J, Marks GC. Moving on from logical frameworks to find the ‘missing middle’ in international development programmes. J Dev Effect. 2019;11(1):89–103.

9. Suchman EA. Principles and practice of evaluative research. New York: Russell Sage Foundation; 1967.

10. Newton-Lewis T, Munar W, Chanturidze T. Performance management in complex adaptive systems: a conceptual framework for health systems. BMJ Global Health. 2021;6(7).

11. Eisenhardt KM. Agency theory: An assessment and review. Academy of Management Review. 1989;14(1):57–74. 10.5465/amr.1989.4279003

12. Mitnick BM. The theory of agency. Public Choice. 1975;24(1):27–42.

13. De Walque D, Kandpal E, Wagstaff A, Friedman J, Piatti-Fünfkirchen M, Sautmann A, et al. Improving effective coverage in health: do financial incentives work? Washington, DC: World Bank Publications; 2022.

14. Davis JH, Schoorman FD, Donaldson L. Toward a stewardship theory of management. Academy of Management Review. 1997;22(1):20–47. 10.5465/amr.1997.9707180258

15. Torfing J, Bentzen TØ. Does stewardship theory provide a viable alternative to control-fixated performance management? Administrative Sciences. 2020;10(4).

16. United Nations Development Programme (UNDP). Human Development Report 2023/24: Reimagining cooperation in a polarized world. New York: United Nations Development Programme; 2024.

17. Bank W. Honduras Poverty Assessment: Toward a Path of Poverty Reduction and Inclusive Growth. Washington, DC: The World Bank; 2024.

18. Carmenate-Milián L, Herrera-Ramos A, Ramos Caceres D, Lagos Ordonez K, Ordoñez TL, Valladares CS. Situation of the health system in Honduras and the new proposed health model. Archivo Medico. 2017;9(4):1–8.

19. Root ED, Zarychta A, Tapia BB, Grillos T, Andersson K, Menken J. Organizations matter in local governance: evidence from health sector decentralization in Honduras. Health Policy and Planning. 2020;35(9):1168–79.

20. Waldron T, Carr T, McMullen L, Westhorp G, Duncan V, Neufeld SM, et al. Development of a program theory for shared decision-making: a realist synthesis. Bmc Health Services Research. 2020;20(1):59.

21. Root ED, Zarychta A, Tapia BB, Grillos T, Andersson K, Menken J. Organizations matter in local governance: evidence from health sector decentralization in Honduras. Health Policy and Planning. 2020;35(9):1168–79. 10.1093/heapol/czaa084

22. Mokdad AH, Colson KE, Zuniga-Brenes P, Rios-Zertuche D, Palmisano EB, Alfaro-Porras E, et al. Salud Mesoamerica 2015 Initiative: design, implementation, and baseline findings. Popul Health Metr. 2015 Feb 7;13:1–16. 10.1186/s12963-015-0034-4

23. Breuer E, Lee L, De Silva M, Lund C. Using theory of change to design and evaluate public health interventions: a systematic review. Implement Science. 2015;11(63). 10.1186/s13012-016-0422-6

24. Mayne J. Theory of change analysis: Building robust theories of change. Can J Program Eval. 2017;32(2).

25. Bhaskar R. A realist theory of science. 1st, editor. London, UK: Routledge; 2010. 240 p. (Classical Texts in Critical Realism).

26. Chen HT. Theory-driven evaluation: Conceptual framework, application and advancement. In: Evaluation von Programmen und Projekten für eine demokratische Kultur. Wiesbaden: Springer; 2012. p. 17–40. 10.1007/978-3-531-19009-9_2

27. Sayer A. Realism and social science. Thousand Oaks: Sage Publications; 2000. 211 p.

28. Rogers PJ. Using programme theory to evaluate complicated and complex aspects of interventions. Evaluation-Us. 2008;14(1):29–48.

29. Leeuw FL. Reconstructing program theories: Methods available and problems to be solved. Am J Eval. 2003;24(1):5–20. 10.1177/109821400302400102

30. Ryan RM, Deci EL. Self-determination theory and the facilitation of intrinsic motivation, social development, and well-being. American Psychologist. 2000;55(1):68–78.

31. Knowles MS. Andragogy in action: applying modern principles of adult learning. San Francisco: Jossey-Bass; 1984.

32. Bandura A. Exercise of human agency through collective efficacy. Curr Dir Psychol Sci. 2000;9(3):75–8.

33. Lave J, Wenger E. Situated learning: Legitimate peripheral participation. Cambridge university press; 1991.

34. Michie S, Stralen MM, West R. The behaviour change wheel: a new method for characterising and designing behaviour change interventions. Implementation science. 2011;6(1):1–12.

35. Nkwake AM. Working with assumptions in international development program evaluation. 2nd ed. Cham, Switzerland: Springer Nature Switzerland; 2020. 212 p.

36. Malterud K, Siersma VD, Guassora AD. Sample size in qualitative interview studies: guided by information power. Qualitative Health Research. 2015;26(13):1753–60. 10.1177/1049732315617444

37. Bernal P, Iriarte E. Coaching to improve quality of primary care-a difference-in difference study in Honduras (Unpublished. Washington, DC: Inter-American Development Bank; 2024.

38. Trochim WM. Outcome pattern matching and program theory. Evaluation and program planning. 1989;12(4):355–66.

39. Franco-Santos M, Lucianetti L, Bourne M. Contemporary performance measurement systems: A review of their consequences and a framework for research. Manage Account Res. 2012;23(2):79–119.

40. Leonard DK. Where are “pockets” of effective agencies likely in weak governance states and why? A propositional inventory. Brighton, UK: Institute for Development Studies; 2008. p. 33. (Working Papers).

41. Gilson L, Grandsoult V. Health systems as human systems: reflexivity, relationships, and resilience in the pursuit of the SDGs. Frontiers in public health. 2025;13:1653839. 10.3389/fpubh.2025.1653839

42. Sheikh K, Gilson L, Agyepong IA, Hanson K, Ssengooba F, Bennett S. Building the field of health policy and systems research: framing the questions. PLoS medicine. 2011;8(8):e1001073. 10.1371/journal.pmed.1001073

43. Hood C. Public Management by Numbers as a Performance-Enhancing Drug: Two Hypotheses. Public Admin Rev. 2012;72:S85–S92.

44. Ebenso B, Mbachu C, Etiaba E, Huss R, Manzano A, Onwujekwe O, et al. Which mechanisms explain the motivation of primary health workers? Insights from the realist evaluation of a maternal and child health programme in Nigeria. BMJ Global Health. 2020;5(8).

45. Lohmann J, Houlfort N, De Allegri M. Crowding out or no crowding out? A Self-Determination Theory approach to health worker motivation in performance-based financing. Social Science & Medicine. 2016 Nov;169:1–8. doi:10.1016/j.socscimed.2016.09.006

46. Vareilles G, Marchal B, Kane S, Petric T, Pictet G, Pommier J. Understanding the motivation and performance of community health volunteers involved in the delivery of health programmes in Kampala, Uganda: a realist evaluation. BMJ Open. 2015;5(11).

47. Wahid SS, Munar W, Das S, Gupta M, Darmstadt GL. Our village is dependent on us. That’s why we can’t leave our work’. Characterizing mechanisms of motivation to perform among Accredited Social Health Activists (ASHA) in Bihar. Health Policy and Planning. 2019;35(1):58–66.

48. Munar W, Wahid SS, Mookherji S, Innocenti C, Curry LA. Team-and individual-level motivation in complex primary care system change: A realist evaluation of the Salud Mesoamerica Initiative in El Salvador. Gates Open Research. 2018;2.

