## Appendix A - Guidelines, Codebook, COREQ for "Coaching for quality improvement under performance-based contracting: a qualitative theory-of-change evaluation in Honduras": Medrvix - Honduras coahing- Appendix A - Guidelines, Codebook, COREQ.pdf

### APPENDIX A – INTERVIEW GUIDELINES, CODEBOOK, COREQ

#### APPENDIX A1- INTERVIEW GUIDELINES

##### 1- MID-LEVEL MANAGERS

Introduction: The purpose of this interview is to help us understand the factors that influence primary health care teams' decisions when implementing national policies for cervical cancer screening and prenatal care.

There are no right or wrong answers.

We want to understand how you approach these topics, including social context, health policies, and the organizational environment of decentralized managers.

In particular, we want to discuss your experiences with the **Salud Mesoamérica Initiative** pilot program regarding training in coaching and quality improvement for cervical cancer screening and prenatal care.

##### SECTION 1

I will begin by asking you a few questions about your professional background.

- What is your current position?
- How long have you held this position?
- How many years of professional experience do you have?
- In the last two years, have you received any training to improve your management skills?

If yes:

- What was the subject of that training?
- When did this training take place?

- What specific elements of that training have helped you the most in carrying out your current responsibilities?

**SECTION 2**

In the following section, we will examine the supervision process for the health units under your charge.

- What types of follow-up and monitoring activities does the central team of this decentralized management unit carry out regarding the health units under its charge?
- How frequently is this follow-up conducted?
- Please describe the most recent follow-up activity you conducted at the health units within your area of influence.
- When did it take place?
- Where did it take place?
- How long did the activity last?
- Who attended?
- What sources of information were used to monitor the performance of the Health Unit?
- Could you give me an example of the type of feedback you provided to the teams under your charge?
- And how does SESAL conduct follow-up and monitoring of the decentralized management unit?
- How frequently does SESAL carry out these follow-up activities? SECTION 3
- In the following section, we will discuss some aspects of the coaching and quality improvement program.

**SECTION 3**

Regarding coaching activities:

- What aspects of the monitoring process that you regularly implement do you find most valuable?
- And for the health teams under your supervision, what do you believe is the added value of the monitoring you conduct?
- How do you think your follow-up activities influence the teams under your supervision? Please provide examples.
- Reflecting on the coaching process as a whole, what were the main lessons learned during your interactions with the health teams?
- Please provide an example related to the quality improvement topics selected by your teams—for instance, cervical cancer or prenatal care.

- And in that specific example, what specific actions did you take following the monitoring activity?
- And which aspects could be improved?

**SECTION 4**

In this final section, we will discuss some of the monitoring actions carried out by SESAL.

- What are the differences between the follow-up activities conducted by the Manager and those carried out by SESAL?
- What are the most valuable aspects of the monitoring conducted by SESAL?
- Which elements could be improved?
- Is there anything else you would like to comment on, or any questions you would like to ask me, before we conclude this interview?

Thank you for your time.

#### 2- PRIMARY CARE TEAM LEADS

Introduction: The purpose of this interview is to help us understand the factors that influence the decisions of primary health care (PHC) teams when implementing national policies for cervical cancer screening and prenatal care.

There are no right or wrong answers.

We want to understand how you approach these topics, including aspects such as the social context, health policies, and the organizational environment of decentralized managers.

In particular, we want to discuss your experiences with the *Salud Mesoamérica Initiative* pilot program regarding training in coaching and quality improvement for cervical cancer screening and prenatal care.

**SECTION 1. We will begin by asking some basic questions about your professional background.**

- In total, how many years have you been practicing?
- What is your current position?
    - How long have you held this position?
    - In the last two years, have you received training to improve your management/administration skills regarding primary care services?

If yes:

- What was the subject of that training?
- When did this training take place?
- What specific elements of that training have helped you the most in carrying out your current responsibilities?

**SECTION 2. Now I will ask you some questions regarding health priorities in Honduras.**

- What are the current priorities of the Ministry of Health regarding the management of cervical cancer (or prenatal care) at the primary care level?
- And what are the priorities regarding this same topic for the decentralized manager?
- Are you familiar with the national standards for cervical cancer screening (or prenatal care)?
  - If yes, can you show me the document containing these standards?
  - Please describe the standard.
  - If no: And where can I find these standards?
- Do you use these standards to guide your daily practice? Do you believe that national guidelines (for the management of cervical cancer—or prenatal care) are useful, or are they a waste of time?
  - Why?
- What are the consequences—positive or negative—of not managing cervical cancer (or prenatal care) in accordance with national guidelines?

**SECTION 3. In the following section, we will discuss the quality improvement project at this health facility.**

- Which improvement project did your team select? (Cervical cancer screening / Prenatal care)
- What were the results? How did your team achieve those results?
- How do these results make you feel? Why do you feel that way?
- What were the most difficult aspects of this project? Why do you think that was the case?
- What were the easiest aspects of the project? Why?
- Give me an example of how your team overcame the challenges encountered during the project's implementation.
- Who supported your team in achieving those results?
- If you could start the project over, what would you need to do to make it work better?
- Why do you think it would work better this time?
- What things should the other members of your quality team do differently?
- What else should the supervisors and coordinators from the decentralized management unit do that they did not do last time?
- What role should SESAL play?

**SECTION 4. In this final section of the interview, we will discuss the capabilities required to improve the management of cervical cancer (or prenatal care).**

- Do you feel that you have the necessary authority to make decisions and act in accordance with your professional knowledge regarding the quality management of cervical cancer screening (or prenatal care)?
- Please explain.
- Do you feel that you have the necessary autonomy to make decisions and act in accordance with your professional knowledge regarding the quality management of cervical cancer screening (or prenatal care)?
- Please explain.
- What type of knowledge, training, or experience should members of the quality improvement team possess to effectively manage cervical cancer screening (prenatal care)?
- What would **you** (not your team) need to do today to improve the way you manage cervical cancer screening (prenatal care)?
- What factors external to your quality team influenced your professional capacity to apply national standards regarding cervical cancer screening (prenatal care)?
- Would any other member of the quality team influence your decision to apply national standards for cervical cancer screening (prenatal care)?
- Who would influence you, and in what way?

**SECTION 5. This is the final section. It differs from the previous ones. It is a mini-survey.**

I am going to read you nine factors known to influence the health care delivery process. Please let me know the extent to which each of them influences patients' care processes (A lot, moderately, a little, or not at all):

- My personal goals influence the care process
- Teams' targets
- Supervisor feedback
- SESAL monitoring
- My salary
- Supervisors' support for my work
- Time availability
- Other daily routine tasks and activities

Can you think of any other influences that you haven't mentioned?

Is there anything else you would like to add?

*Thank you for your participation!*

##### 3- MANAGERS AT SESAL REGIONAL OFFICES

**SECTION 1. We will begin by asking you some basic questions about your professional background.**

- What is your current position?
- How long have you held this position?
- What is the most significant leadership position you have held throughout your career?
- Have you received any training in health services management? If so:
- What did that training cover?
- When did this training take place?
- Which aspects of that training have helped you the most in your role as a SESAL manager?

**SECTION 2. In the following section, we will examine the monitoring process for decentralized managers.**

- What types of monitoring or follow-up activities does SESAL conduct with decentralized managers?
- How frequently do you carry out these activities?
- And, in turn, how do decentralized health operators monitor the performance of their own health units?
- In what ways do you believe monitoring activities influence decentralized managers?
- Please give me an example.
- And, are there any undesirable effects? Please give me examples.
- In your view, what are the most valuable aspects of this monitoring/follow-up process?
- Why?
- Which elements of the monitoring process do you think could be improved?
- What benefits would these improvements yield regarding the quality of care?

**SECTION 3. We will now discuss the coaching and quality-of-care program.**

- Please describe the program's positive aspects for the participating health units.
- And for the decentralized manager?
- And for the SESAL health regions?
- And for the central level of SESAL?

- Which aspects of the model could be improved?
- What benefits would these adjustments bring?

Is there anything else you would like to comment on, or any questions you would like to ask me, before we conclude this interview?

*Thank you for your time.*

#### APPENDIX A2 – CODEBOOK

| Codes | Comment |
|---|---|
| A.0 Capability | **Definition:** Capability refers to the individual's psychological and physical capacity to engage in the activity concerned. It includes having the necessary knowledge and skills. |
| A1. Psychological Capability | **Definition:** The team's knowledge, cognitive abilities, and problem-solving skills needed to make informed decisions, adapt to context, and deliver care. |
| A.1.1 Knowledge of MOH priorities | **Definition:** Awareness of the current priorities of the Secretary of Health (MOH) for cervical cancer screening or prenatal care.<br>**When to use?** Apply when the respondent outlines what they believe are the MOH's key focus areas |
| A.1.2 Knowledge of Operator's priorities | **Definition:** Awareness of the priorities of the decentralized manager for cervical cancer screening or prenatal care.<br>**When to use?** Apply when the respondent discusses the manager's priorities, which may differ from MOH. |
| A.1.3 Knowledge of national standards of care | **Definition:** Familiarity with the national standards for cervical cancer screening or prenatal care.<br>**When to use?** Apply when standards are mentioned, including if the respondent can produce the standards document, describe the standards, and whether they routinely apply them in practice |
| A.1.4 Knowledge & skills needed for QI project | **Definition:** The knowledge, training and experience the respondent feels are needed by quality improvement team members to effectively manage cervical cancer screening or prenatal care.<br>**When to use?** Apply when the respondent outlines key competencies required for implementing the chosen quality improvement project. |
| A.2 Physical Capability | **Definition:** The team's physical skills to perform tasks efficiently, such as using medical equipment and medications, documenting patient records, and delivering care. |
| A.2.1 Years practicing | **Definition:** Total number of years the respondent has been practicing in their field.<br>**When to use?** Apply this code to respondents' answers about their total years of work experience. |

| | |
|---|---|
| A.2.2 Training | **Definition:** Any mention of training the respondent has received in the past 2 years.<br>**When to use?** Apply when the respondent discusses recent relevant training they have completed. |
| B.0 Opportunity | **Definition:** The physical and social factors that enable OR prevent the behavior from occurring. It refers to all the factors that lie outside the individual that make the behavior possible or prompt it. |
| B.1 Physical Opportunity | **Definition:** Access to the necessary equipment, resources, and infrastructure to effectively deliver care. This includes well-stocked medical supplies, functional technology, and efficient workflows, among others. |
| B.1.1 Time availability | **Definition:** The degree of influence that time availability has on the care process, based on respondent's experience.<br>**When to use?** Apply to statements describing having time (or not) to complete QI-related activities. |
| B.1.2 Competing responsibilities | **Definition:** The degree of influence that other routine activities the respondent is responsible for have on the QI process.<br>**When to use?** Apply to statements describing having responsibilities (or not having them) that compete with the QI-related activities. |
| B.2 Social Opportunity | **Definition:** Collaborative and supportive work environment that enables the team to communicate effectively, share knowledge, and work together towards common goals. Spaces or events promoting a culture of teamwork, adaptation, learning, and continuous improvement. |
| B.2.1 Supervisor feedback | **Definition:** The degree of influence that feedback received from the supervisor has on the care process.<br>**When to use?** Apply to descriptions of team's supervisor feedback, or lack thereof. |
| B.2.2 Supervisor support | **Definition:** The degree of influence that support received from the supervisor has on the care process.<br>**When to use?** Apply to respondent's descriptions of supervisor's support for their work or that of the team's or lack thereof. |
| B.2.3 MOH monitoring | **Definition:** The degree of influence that SESAL monitoring has on the care process.**When to use?** Apply to respondent's descriptions of SESAL monitor of Operator's work. It may refer to monitoring conducted by the central and regional offices of the MOH. |

| | |
|---|---|
| B.2.4 Team members' influence | **Definition:** Ways in which other members of the QI team influence the respondent's ability or decision to implement standards of care for CCS or ANC.<br>**When to use?** Apply when the respondent discusses how team members impact each other's actions. |
| B.2.5 Other sources of support | **Definition:** Any individual or group that provided support to the team to help achieve results.<br>**When to use?** Apply when the respondent mentions receiving support to accomplish their efforts from stakeholders other than the MOH or the Operator (e.g., SMI and others). |
| C.0 Motivation | **Definition:** Motivation refers to all the brain processes that energize and direct behavior. It includes habitual processes, emotional responding, as well as analytical decision-making.<br>**Comment:** This is not a synonym with **work motivation.** |
| C.1 Reflective Motivation | **Definition:** The team's ability to consider their actions, evaluate the evidence, and make conscious decisions that align with the initiative's goals and their own values. For example: Teams evaluating the data in their QI practice. |
| C.1.1 Personal goals | **Definition:** The degree of influence the respondent's personal goals have on the care process.<br>**When to use?** Apply to quotes describing experiences in which respondents describe how personal goals inform their use or nonuse of standards of care.<br>**Comment:** Respondents describe how the care process fits/aligns (or not) with their personal goals |
| C.1.2 Team goals | **Definition:** The degree of influence the improvement team's goals have on the care process.<br>**When to use?** Apply to quotes describing experiences in which respondents describe how team goals inform their use or nonuse of standards of care.<br>**Comment:** Same as above but more related to Locke's theory**.** |
| C.1.3 Beliefs about consequences | **Definition:** Any positive or negative consequences of managing cervical cancer or prenatal care according to national standards of care.<br>**When to use?** Apply when a respondent describes implications from adhering to or deviating from standards of care.<br>**Example:** When we follow the standards I believe it leads to better care for our patients |

| | |
|---|---|
| C.1.4 Feelings about results | **Definition:** The respondent's emotional reaction to and reflections on the results achieved through the QI project.<br>**When to use?** Apply when feelings/emotions about the outcomes are described.<br>**Example:** I feel proud of how we now get results completed quicker |
| C.2 Automatic Motivation | **Definition:** Individual and team's habits, instincts, and routines that drive them to perform their duties and provide high-quality care without having to consciously think about it.<br>**Examples:** Action plans, and other routines/systems such as driving the samples at certain times and hours (scheduling). |
| **D.0 Context** | **Definition:** The institutional, organizational, spatial, and temporal settings in which interventions are implemented (1), as well as the interpersonal relations within and across levels of the health system, the individual and group actors targeted by the intervention, and the external environment (2).<br>• Incentives<br>• External incentives (i.e. non-salary incentives, in-kind as well as financial other than salary) on the care process<br>• Other external incentives (i.e., non-salary incentives, in-kind as well as financial other than salary) on the care process. |

| | |
|---|---|
| D.1 Institutional context | **Definition:** Refers to salient conditions at the facility, operator, or higher institutional level that may favor or disfavor implementation and outcomes of the intervention.<br>**When to use?** Apply to descriptions of organizational conditions at the level of the facility, operator, or the MOH units involved. Respondents may refer to specific aspects of the physical and social environment that may influence (positively or negatively) the expected outcomes. Such components may include:<br>• The history and antecedents of supervision and management practices.<br>• The quality of existing health system networks and communications as it pertains to the formal and informal social interactions connecting facilities and operators in the health network.<br>• Leadership support.<br>• **Availability (e.g., supplies, equipment, time, knowledge) and flow of resources (financing, data/information, etc.)**<br>• (Workplace) Incentives including salary<br>**• Policies, rules and regulations by the SESAL (i.e., pay for performance, contracting operators, etc.) as well as by the Operators (internal strategies and rules such as using checklists to implement providers' supervision).** |
| D.2. Inner setting | **Definition:** Refers to the organizational environment within the operators and teams that may favor or disfavor implementation and outcomes of the intervention.<br>**When to use?** Apply this code when respondents refer to interpersonal relationships, communication, networking, and learning, among others, within the operators and teams. This code may also include statements related to collective norms and values. |
| E.0 Behavioral outcomes | **Definition:** Refers to positive or negative changes produced by an intervention – directly or indirectly, intended, or unintended – at the individual, team, or organizational level. |
| E.1 Individual-level behavioral outcomes | **Definition:** Behaviors that were expected in the design at the level of Supervisors and/or Coordinators, and QI team members as well as unexpected or unplanned behaviors. |
| E.1.1 (As an example) Use of national standards of care | **Definition:** Extent to which the respondent uses national standards to guide their daily practice.<br>**When to use?** Apply when the respondent indicates if they routinely apply national standards in service delivery. |

| | |
|---|---|
| E.1.2 New QI skills | **Definition:** Developed basic capacities at the managerial (operator) and facility levels (PHC teams) to conduct the steps and activities of PDSA cycles to improve quality care.<br>**When to use?** Apply when respondents describe hard skills acquired to carry out QI PDSA cycles, such as measurement, monitoring, and information usage. |
| E.1.3 Soft skills for coaching | **Definition:** Developed capacities to lead, support, and empower PHC teams to undertake PDSA cycles to improve quality care.<br>**When to use?** Apply when respondents describe soft skills developed such as leadership, supportive supervision, and communication. |
| E.1.4 Job satisfaction | **Definition:** Extent to which respondents express feeling valued, important, or respected (3) AR.<br>**When to use?** Apply when the respondent refers to emotional reactions and feelings about work |
| E.1.5 Organizational Commitment | **Definition:** Increase or decrease in willingness to exert and maintain an effort towards organizational goals (3).<br>**When to use?** The individual is committed to fulfilling their Role (Follows CFIR characteristics subdomain for motivation) |
| E.2 Interpersonal, team, and organizational level outcomes | **Definition:** Organizational behaviors that can plausibly occur at the level of teams, operators, and the MOH units involved. Examples include coordination across organizations (e.g., SESAL and operators), inter-team collaboration, diffusion & dissemination effects.<br>**When to Use:** If teams have new capacity (i.e., C+M+O), what specific practices (also networks, relations, coalitions, changes in power dynamics, etc.) they are doing together as team they were not doing before? The same for the Operator and possibly but unlikely from the MOH |
| E.3 Unexpected, negative, and emergent outcomes | **Definition:** Behaviors that were unplanned, negative, or emergent (resulting from the interactions between levels of analysis or plausibly triggered by contextual conditions). This can be a catch-all, broad category. |
| **F.0. Intervention** | **Definition:** Interventions designed to bring about changes in healthcare organizations, the behavior of healthcare professionals, or the use of health services by healthcare recipients (4). |

| | |
|---|---|
| F.1.1 In-person training by external tutors | **Definition:** Refers to training for regional supervisors and operator coordinators on soft skills such as teambuilding, coaching, communications and leadership and hard skills such as measurement, problem-solving, and applying the Model of Improvement (including PDSA cycles).<br>**When to use?** Apply to descriptions of the training received from external trainers. |
| F.1.2 In-person monitoring and mentoring at worksite | **Definition:** Refers to monitoring and mentoring conducted by external trainers.<br>**When to use?** Apply to descriptions of the onsite monitoring and coaching activities conducted by external trainers. |
| F.1.3 1-week QI training and QI collaboratives | **Definition:** Refers to regional supervisors and operator coordinators training PHC teams on QI concepts to enable teams to develop QI projects. Supervisors and coordinators also supervised and coached PHC teams during QI project development and implementation.<br>**When to use?** Apply to descriptions of training sessions with PHC teams and onsite and remote coaching to monitor their progress on QI projects. |
| F.1.4.1 QI training of SESAL regional directors | **Definition:** Refers to training for SESAL regional directors on QI (on soft skills such as teambuilding, coaching, communications and leadership and hard skills such as measurement, problem-solving, and applying the Model of Improvement (including PDSA cycles).<br>**When to use?** Apply to descriptions of the training received from external trainers. |
| **G.1 Inductive Codes** | **Use this for codes that emerge during the analysis not otherwise captured.** |
| D.3 External Context | **Definition:** Captures community social norms, environment |
| E.1.6 Communication | **Definition:** Capture interpersonal dialogue between hierarchies (supervisors to health teams)<br>**When to Use?** Apply when discussing shifting from punitive approaches to more supportive and collaborative dialogue to problem solve together |

| | |
|---|---|
| E.1.7 Change in attitude | **Definition:** Capture the changes in perspectives of shifts in the power dynamic between hierarchies (supervisors to health teams) to a collaborative environment (idea of accompaniment)<br>**When to Use?** Apply when discussing shifting from punitive approaches to more supportive and collaborative dialogue to problem solve together |
| B.2.6 Supervisor visits | **Definition:** Routine visits by supervisors to health teams.<br>**When to Use?** Apply when discussing routine visits to health teams by supervisors. |

#### Citations


1. Coldwell M. Reconsidering context: Six underlying features of context to improve learning from evaluation. Evaluation. 2019;25(1):99–117. https://doi.org/10.1177/1356389018803234
2. Pawson R. The science of evaluation: a realist manifesto. London: Sage Publishing; 2013. 216 p.
3. Aranda LE, Arif Z, Innocenti C, Wahid SS, Frehywot S, Munar W. Characterizing the implementation of performance management interventions in a primary health care system: a case study of the Salud Mesoamerica Initiative in El Salvador. Health Policy Plan. 2023; czad020. https://doi.org/10.1093/heapol/czad020
4. EPOC. Effective Practice and Organisation of Care (EPOC). EPOC taxonomy [Internet]. Cochrane Collaboration. Norwegian Satellite; 2015 [cited 2019 Oct 17]. Available from: https://epoc.cochrane.org/epoc-taxonomy

#### A3. COREQ Checklist

| No. | Domain / Item | Location in manuscript |
|---|---|---|
| **DOMAIN 1: Research Team and Reflexivity** | | |
| **Personal Characteristics** | | |
| **1** | Interviewer / facilitator | p. 5 |
| **2** | Credentials | p. 5 |
| **3** | Occupation | p. 5 |
| **4** | Gender | p. 5 |
| **5** | Experience and training | p. 5 |
| **Relationship with Participants** | | |
| **6** | Relationship established | p. 5 |
| **7** | Participant knowledge of the interviewer | p. 5 |
| **8** | Interviewer characteristics | p. 5 and 6 Table 1) |
| **DOMAIN 2: Study Design** | | |
| **Theoretical Framework** | | |
| **9** | Methodological orientation and theory | p. 1–4 |
| **Participant Selection** | | |
| **10** | Sampling | p. 5 |
| **11** | Method of approach | p. 5 |
| **12** | Sample size | p. 5 |
| **13** | Non-participation | p. 5 |
| **Setting** | | |

| | | |
|---|---|---|
| **14** | Setting of data collection | p. 5 |
| **15** | Presence of non-participants | p. 5 |
| **16** | Description of sample | Table 1 (p. 6) |
| **Data Collection** | | |
| **17** | Interview guide | p. 6 and Appendix A |
| **18** | Repeat interviews | p. 5 |
| **19** | Audio / visual recording | p. 5 |
| **20** | Field notes | p. 5 |
| **21** | Duration | p. 5 |
| **22** | Data saturation | Not applicable |
| **23** | Transcripts returned | p. 6 |
| **DOMAIN 3: Analysis and Findings** | | |
| **Data Analysis** | | |
| **24** | Number of data coders | p. 5 |
| **25** | Description of the coding tree | Appendix A |
| **26** | Derivation of themes | p. 5; Appendix B |
| **27** | Software | p. 5 |
| **28** | Participant checking | p. 6 |
| **Reporting** | | |
| **29** | Quotations presented | p. 7–9; Appendix B |
| **30** | Data and findings consistent | Table 2 (p. 12) |
| **31** | Clarity of major themes | p. 7–9; Table 2 |
| **32** | Clarity of minor themes | Appendix B |
