## APPENDIX B: Indicative quotes for "Coaching for quality improvement under performance-based contracting: a qualitative theory-of-change evaluation in Honduras": Medrvix - Honduras Coaching - Appendix B.pdf

### Theme: Capability

**Managers**. The findings show managers' capability to be a multifaceted construct primarily centred on psychological, knowledge-based, and skill-based dimensions. Shared viewpoints consistently highlight psychological resources—often built through coaching—as foundational to confidence, motivation, and the creation of supportive team environments. Furthermore, explicit knowledge of formal frameworks (MOH priorities, national standards) and a comprehensive QI skill set are universally recognized as core components of professional capability. However, unique perspectives add critical nuance, showing that capability is also sustained by the meaningful impact of work, involves a critical awareness of systemic disconnects, and notably excludes physical dimensions in practical discourse. This analysis demonstrates that capability is not a monolithic trait but an integrated, context-dependent combination of internal resources, applied knowledge, and practical skills, with its expression and development shaped by both structured support and the experiential realities of the work environment.

**Providers**. The synthesis of Providers' Capability shows a complex and multi-layered construct within the Honduran primary healthcare context. A strong shared perspective defines capability as the possession of specific, often procedural, knowledge—of ministerial and operator priorities, clinical standards, and quality improvement (QI) methodologies—coupled with the skills to apply this knowledge through measurement, teamwork, and adaptive problem-solving. Distinct physical resource constraints are also recognized as a shared barrier to delivering specific services, such as cervical cancer screening. However, unique viewpoints significantly deepen this understanding. They reveal critical gaps: a complete absence of explicit discussion of psychological factors such as confidence; sophisticated critiques of implementation priorities and inadequate equipment design; a foundational lack of administrative training among clinical leaders; and pronounced variability in technical QI skills within teams. Finally, the silence on Physical Capability in most transcripts and the transformative impact of effective training on team agency highlight that capability is not a uniform set of attributes but a dynamic, context-dependent combination of knowledge, skills, material resources, and unspoken psychological readiness, heavily influenced by the quality of professional development and support structures.

### Indicative Quotes

| Managers (OP-) | PHC Team Leads<br>(HF-) | Regional SESAL Directors (DIR-) |
|---|---|---|
| OP-03 —"And as a supervisor, I sometimes feel like I don't know anything..." | HF-04 — "For example, yesterday, the nurse told me, Doctor if you are going to do cytology, there are no gloves." | DIR-01 — "So, creating awareness, that was one of the things I did when I had that administrative knowledge and then |

| Managers (OP-) | PHC Team Leads (HF-) | Regional SESAL Directors (DIR-) |
|---|---|---|
| OP-12 — "We have also learned to shape our attitudes, emotions, and character in terms of handling staff, which is very important. From not accusing or pointing fingers but seeing first what is positive and, in some way or another, then what is not good." | HF-02 — "When the intervention was made in cervical cancer screening, we implemented several formats for weekly evaluations or monitoring, which made it much easier to identify where the problem was. And it worked very well for us." | the will of the team, the capacity of the team to replicate it to the rest that did not directly receive the diploma and see the results." |
| OP-07 — "Yes, it is worth it... But when this was applied, we saw the difference in the uptake of cytology there was even one of the health centers... which detected two women with cervical cancer... who had never had a cytology." | HF-01 — "Well, it would be important to have a general training for all the staff, because as a physician you already know about cervical cancer and cytology. About what the results could be, but a health promoter does not. So, yes, it would be good to have training for all the staff on continuous improvement." | |

| Managers (OP-) | PHC Team Leads (HF-) | Regional SESAL Directors (DIR-) |
|---|---|---|
| OP-12 — "The most valuable thing about monitoring is the evaluation of standards. We, as decentralized health operators, must adhere to the standards, and that's the most valuable thing." | — | |
| OP-05 — "it changed our lives professionally because it removed a chip that we had" | — | |

#### Theme: Opportunities

**Mid-Level Managers.** The findings about opportunity in the supervisory context show a deeply constrained yet complex and dynamic operational environment. The opportunity for effective quality improvement work is fundamentally shaped by severe and pervasive physical barriers, particularly the scarcity of time and material resources, which act as a baseline constraint. This is compounded by a hierarchical and often punitive system of external monitoring that can undermine local efforts. However, the analysis also uncovers powerful social and strategic counterforces that actively create opportunity. A transformative shift in supervisory philosophy towards collaboration and capacity-building has created a vital social opportunity for team empowerment. Furthermore, participants demonstrate resilience and adaptability by strategically leveraging external partnerships, navigating local politics, and even engineering physical opportunities through resource reallocation. The opportunity landscape is therefore not static; it is a site of constant negotiation where structural limitations are met with proactive, relationship-based strategies to enable improvement work.

**Providers**. Overall, the findings reveal providers' opportunity as a dynamic, multi-layered construct that encompasses both environmental conditions and agentive actions. Shared viewpoints converge on the critical opportunities created internally through cohesive teamwork, hands-on supervisory partnerships, and proactive logistical and social engineering by frontline staff. These demonstrate that opportunity is often cultivated from within through collaboration, optimized processes, and strategic communication. Unique perspectives add nuance, showing that opportunity can also be sought through advocacy for systemic policy change, yet it can be simultaneously constrained by bureaucratic red tape. Furthermore, an exceptional case reveals that opportunity may be seized through informal political channels to meet acute patient needs. The collective evidence underscores that while structural and systemic barriers are pervasive, healthcare teams and leaders actively identify, create, and leverage social, logistical, and political opportunities to drive improvement, though their success is often contingent on navigating a complex landscape of support and constraint.

### Indicative Quotes

| Managers (OP-) | PHC Team Medical Leads (HF-) | Regional SESAL Directors (DIR-) |
|---|---|---|
| OP-03 — "We used to do a monthly one, but they are not complete supervisions because due to the time factor of a full day in a facility, one feels that one cannot carry it out 100." | HF-03 — "Something my team did. Well. I think working all on the same goal. Everybody committed. Everybody changed their attitude. And with that, we had already won the friendship." | DIR-01 — "the will of the team, the capacity of the team to replicate" |
| OP-08 — "at least 400 cytology... is quite impossible to do with the resources we have" | HF-02 — "To date, we have not worked more than the work schedule, and we have managed to get through all the activities." | DIR-01 — "making higher authorities see that we have to improve these monitoring instruments" |
| OP-05 — "shift from decentralized to centralized has made management processes slower and more cumbersome... This situation creates delays of three to four months" | HF-01 — "started sending the samples faster every Friday" | DIR-01 — "the regulations are about to come out in about three, four months" |

| Managers (OP-) | PHC Team Medical Leads (HF-) | Regional SESAL Directors (DIR-) |
|---|---|---|
| OP-02 — "I feel that... my team sees me as a co-worker; we are horizontal... all we are really looking in the same direction" | | |
| OP-01 — "each member of the health region... no longer came with the attitude of only seeing the bad. They also saw the opportunities for improvement" | | |

### Theme: Motivation

**Managers**. Overall, the findings on managers' motivation show a multifaceted construct central to the experiences of supervisors and medical coordinators. The shared viewpoints converge on a deep professional transformation, where participants consciously shifted their identity towards coaching, grounded their intentions in patient-centred duty and purpose, and set goals focused on team empowerment and sustainable collaboration. These changes are sustained by strong beliefs in the positive consequences of new methods and in their own enhanced capabilities. However, this positive reflective drive is not universal; unique perspectives illustrate its fragility when confronted with demotivating external factors such as inconsistent oversight or unrealistic targets. Furthermore, the concept itself was not explicitly engaged with by all participants, indicating variability in how motivations are articulated versus analysed. In essence, reflective motivation emerges as a dynamic, context-sensitive driver of professional change, powerful when supported by positive beliefs and a conducive environment, yet vulnerable to external pressures that can disrupt the conscious commitment to improvement.

**Providers**. Overall, the findings show providers' motivation as a multidimensional and dynamic construct, shaped by a core interplay between internally-driven reflective elements and more immediate automatic responses. Shared viewpoints show that motivation is fundamentally anchored in the intrinsic satisfaction of providing life-saving, high-quality care and in the conscious pursuit of personal and team goals, which are seen as essential for direction and performance. This drive is powerfully reinforced by positive beliefs in the consequences of their work and feelings of satisfaction from tangible results, yet it is equally vulnerable to reflective assessments of systemic barriers and feelings of frustration from stagnation. Unique perspectives add critical nuance, showing that motivation is not static but can be significantly reshaped by catalytic events such as external validation, can be trivialized by unambitious targets, and that achieving uniform motivation across a team is challenged by skill disparities. The analysis highlights that sustaining high motivation in this context requires not only nurturing internal purpose and collaborative processes but also proactively managing external demotivators and creating environments that validate effort and enable habitual, patient-centred engagement.

**Indicative Quotes**

| **Managers (OP-)** | **PHC Team Medical Leads (HF-)** | **Regional SESAL Directors (DIR-)** |
|---|---|---|
| OP-02 — "I have had better results now than before because before, I achieved everything, perhaps in an angry way, as we say, or as we say empirically, in a rough way. And, really, the results are not the same as when you have a preparation, and the results you see are better." | HF-05 — "This is a very demotivating factor for everyone." | DIR-02 — "You are just going to be there and give the minimum you can give" |
| OP-04 — "I think that they learned, as I tell you, for me, measuring what you do is very important, and I think this is something that they learned and learning to analyze the results of what they are getting." | HF-06 — "Well, I will tell you that most of us have a contract signed until June of this year... I feel a little bit uncertain about the situation." | DIR-01 — "now with the change of government, there has been a little bit of a split in these agreements" |
| | HF-04 — "Right now, our cytology targets are low. It's only 13 cytologies per month which is very low! So, since the | |

| Managers (OP-) | PHC Team Medical Leads (HF-) | Regional SESAL Directors (DIR-) |
|---|---|---|
| | goal was low, it was not very difficult to achieve." | |
| | HF-06 — "From the moment of formulating an indicator, it may be easy for me, but maybe not for my colleague, so I have to explain it to them." | |
| | HF-02 — "We really feel satisfied because we have the opportunity to save a person's life and not only in screening, but this part of quality has helped us in many processes that are provided in the health facility." | |
| OP-05 — "Before, there was monitoring under the management agreements with which the operators worked. But when we changed the model, we were no longer a decentralized model; we were a centralized model." | HF-02 — "Yes, yes, we work based on national standards and protocols already established by the Ministry of Health." | |
| OP-02 — "…the plans, so to speak, have no follow-up so far. Okay, so, for example, in the region that we are basically linked to right now, there is no strict follow-up, so to speak, to an improvement plan. So we are a little weak in that part, I would say, because we do not have a follow-up as it should be…" | HF-06 — "Even though it's decentralized now, it has always worked with the same regulations." | |
| OP-12 — "The truth is that the procedures were not as cumbersome as before when they were centralized. Health is now decentralized, and the funds are managed here, and the attention is more expeditious. Problems are solved more quickly, of course. More agile." | HF-04 — "So, in that sense I have no authority and no autonomy." | |

| Managers (OP-) | PHC Team Medical Leads (HF-) | Regional SESAL Directors (DIR-) |
|---|---|---|
| OP-03 — "For example, when errors are found during the monitoring, they should explain to us how to do it differently. Excellent. How do we do it correctly because we are making mistakes, but they do not explain the correct way to do it. Then we will keep doing the same thing, and obviously, we will keep postponing it or losing the monitoring points because we don't know how to do it correctly." | HF-05 — "The economic factor has always been a problem. Many times, we had to buy some supplies out of my pocket..."<br><br>HF-03 — "I should comment that the culture in which we are working is the Lenca Peoples culture. So their culture is well rooted with their beliefs and customs." | |

### Theme: Outcomes

**Managers**. Overall, the behavioural outcomes reveal a complex and often contradictory landscape. Direct improvements in providers' patient care behaviours—specifically the adoption and adherence to national standards for cervical cancer and antenatal care—are explicitly identified as the program's ultimate goal. This intent is substantiated by consistent reports of the application of new technical QI skills and structured project management, which aim to systematize and improve care. The most profound and widely shared behavioural outcomes, however, are found in the interpersonal and leadership domain. Participants demonstrated a marked shift from authoritarian to collaborative, empowering supervisory practices, characterized by improved communication, active listening, and co-creative team leadership. These new behaviors engendered greater team satisfaction, commitment, and a reconceptualization of supervision from punitive to supportive.

However, these positive internal behavioural shifts exist in constant tension with powerful external behavioural drivers, namely a rigid, top-down performance monitoring system that enforces outdated national standards through punitive audits and unrealistic targets. This system is reported to inflict significant negative outcomes, including profound demoralization, feelings of betrayal, and a reactive sense of futility among frontline health workers. Therefore, while the intervention successfully cultivated a new set of empowering and relational supervisory behaviours, their sustainability and broader impact on health outcomes are critically undermined by persistent, demotivating behaviours enforced by the larger institutional and external regulatory environment.

**Providers**. Overall, the analysis reveals that behavioural outcomes in primary healthcare settings are shaped by a dynamic interplay between positive developments and systemic challenges. Shared viewpoints highlight a common trajectory in which Quality Improvement (QI) interventions and coaching foster significant positive behaviours, including the proactive use of standards, the acquisition of systematic problem-solving skills, enhanced team communication and collaboration, increased job satisfaction rooted in meaningful impact, and resilient local team commitment. These outcomes collectively contribute to improved efficiency, service delivery, and a shift towards empathetic, patient-centred care. However, these positive developments are consistently and powerfully challenged by shared negative behavioural outcomes stemming from systemic failures, such as demoralization due to resource constraints and inadequate support, compromised adherence due to overwhelming workloads, and communication breakdowns. Unique viewpoints provide critical nuance, revealing extreme cases of toxic interpersonal dynamics that sabotage teamwork, deliberate non-compliance driven by safety concerns, the profound fragility of positive outcomes amid high staff turnover, and the lamentable ultimate behavioural consequence of plausible patient harm resulting from system collapse. The synthesis underscores that sustainable positive behavioural change requires not only effective local interventions but also a supportive and stable organizational environment to mitigate the demotivating and destructive forces inherent in under-resourced systems.

### Indicative Quotes - Outcomes

| Managers (OP-) | PHC Team Medical Directors (HF-) | Regional SESAL Directors (DIR-) |
|---|---|---|
| OP-01 — "I am the boss. I do this. And I want this and that" | HF-04 — "I am not going to change any files; I am not going to touch any of them... I don't want to start inventing" | DIR-02 — "people work a little bit based on a goal... when you have a monitoring system, the person is aware of what is going to be evaluated." |
| OP-02 — "The communication is better with the staff. They understand them better. The communication is reciprocal, and also then at the end that gives us better results." | HF-02 — "it was really less burden" | DIR-02 — "They influence positively. Definitely at the beginning they didn't like it very much because it was more, more intense and more work. Yeah, but in that sense it's quite positive." |
| OP-10 — "Well, the monitoring that SESAL does for us, let's say, they monitor compliance with monitoring indicators ... every three months. They choose a health facility at random. And that is punitive for us. If we don't have something, they take away points." | — | DIR-01 — "What we have seen as a weakness is that these monitoring instruments fall short. They are not accurate; they do not cover as much as they should." |
| OP-07 — "When he gave us the replication of this coaching, at the beginning, he only called the doctors, the managers, the bosses. I told him, Look, you went there, you received all the information, you retained some of it, you replicated it to me, and I am going to replicate even less to my team. So, when there were meetings, he should have brought us all together so that we could all understand what he was giving us." | — | DIR-01 — "I do not say to the operator solve it. I say when are we going to solve it, and we go here with the regional team to support them because for me they are not operators, that is, for me, we are all the same team..." |
| OP-08 — "Well, what I think is that I feel that soft skills are a fundamental tool because if people don't have the attitude, they are not motivated to do the job as it should be." | — | DIR-02 — "I will go back to a little bit stricter monitoring because they should go back to the monitoring as it was before" |
